# A double-blind, randomised, placebo-controlled trial to evaluate the effectiveness of late gestation oral melatonin supplementation in reducing induction of labour rates in nulliparous women. The MyTIME study protocol

**DOI:** 10.1101/2024.08.20.24312284

**Authors:** Z. Bradfield, S. White, M. Davies-Tuck, M. Sharp, J. Warland, E. Callander, L. Kuliukas, M. Rose, A. Pettitt, K. Ekin, D. Doherty, J. Keelan

**Affiliations:** Curtin University, Women and Newborn Health Service; Division of Obstetrics and Gynaecology, Medical School, The University of Western Australia; Hudson Institute of Medical Research and Dept O and G, Monash University; Perth Children’s Hospital; University of Adelaide; University of Technology Sydney; Curtin University Bentley Campus: Curtin University; Curtin University; University of Western Australia

## Abstract

**Introduction:** Around the world, rates of induction of labour (IOL) amongst nulliparous mothers have increased in the last 10 years. In Australia, rates have increased over the last decade by 43%, from 32% to 46%. There is growing concern about the rapid rise in IOL before 41 weeks for nulliparous women without medical complications because of the associated increased rates of caesarean section, reduced satisfaction with birth, and birth trauma. Melatonin potentiates the action of oxytocin and may promote the spontaneous onset of labour; therefore, we will test the hypothesis that exogenous melatonin supplementation in late pregnancy will reduce the rate of labour induction by 30% or more.

**Methods and analyses:** This is a double-blind, randomised, placebo-controlled trial in nulliparous pregnant women to reduce IOL rates. We will randomise 530 women to receive either 3 mg oral melatonin or placebo daily from 39^+0^ weeks gestation until they give birth. The primary endpoint will be IOL rate after 39 weeks. Secondary endpoints will include: interval between administration of trial medication and birth; a range of maternal and neonatal outcomes, including birth outcomes; breastfeeding on discharge, at 10 days and at 2 months; maternal satisfaction; child developmental outcomes at 2 months of age; and cost-effectiveness of melatonin compared with standard care. All data will be analysed by intention to treat.

**Ethics and dissemination:** The study is approved by the Women and Newborn Health Service (WNHS) Human Research Ethics Committee (HREC) (RGS0000006283).

Trial findings will be disseminated through conference presentations and peer reviewed publications.

**Trial registration number:** The trial has been prospectively registered on the Australian New Zealand Clinical Trials Registry as ACTRN12623000502639

**Strengths and Limitations:** 

- This is the first registered randomised placebo-controlled clinical trial investigating the effect of melatonin to improve the onset of spontaneous labour from 39 weeks gestation and therefore, reduce the rates of women requiring IOL. Both participants and clinicians will be blinded to intervention allocation, ensuring decisions regarding induction of labour cannot be influenced by the allocation.
- Exploration of child developmental outcomes is a strength, not previously undertaken in trials including melatonin supplementation in pregnancy.
- Sub study findings from participants with Gestational Diabetes Mellitus (GDM) will provide novel pilot data regarding the potential role for melatonin in blood glucose control.
- A limitation of our trial is that is conducted within one health service area which may limit generalisability; however, this area is large, encompassing 4 separate sites, various models of care and all capability levels of maternity care (primary through to quaternary) minimising the impact of this selection bias.
- Singular health service governance over such a large variety of settings supports protocol implementation and compliance, reducing variability and improving trial quality.

## INTRODUCTION

There is growing concern around the world about the rapid rise in IOL before 41 weeks, in the absence of maternal or fetal complications, because of the risk of iatrogenic harm to the mother and baby (1-6). Obstetric intervention in late pregnancy has inexorably risen over recent decades, more than is accounted for by changing population complexity and risk factors; this has occurred without a demonstrated reduction in stillbirth, and with a clear increase in early-term birth (37–38 weeks) which is known to be harmful to subsequent childhood neurodevelopment (7). In particular, there is concern about the increased incidence of caesarean section, Neonatal Intensive Care Unit (NICU) admission, and adverse longer-term neurodevelopmental outcomes for children born prior to 39 weeks or after 41 weeks (2-6, 8). Other perinatal morbidity indicators are also rising, including episiotomy, postpartum haemorrhage, maternal sepsis, and, maternal and neonatal birth trauma (4, 9-12).

The World Health Organization (WHO) guidelines indicate that, for those women whose pregnancies are not complicated, IOL prior to 41 weeks is not recommended (13). For pregnant women without medical complication, the most common reason for IOL is prolonged pregnancy beyond 42 completed weeks (9). Recent evidence from studies around the world reveal that amongst women, the broad acceptability of IOL is low (2, 14) and may increase the risk of traumatic stress (15). There is limited evidence regarding the long-term perinatal outcomes of induction of labour. One Australian retrospective cohort study showed increased incidence of hospital admission for ear, nose, throat, respiratory complaints and sepsis up to 16 years of age (4). A retrospective cohort study in the Netherlands revealed an association between induction of labour and reduced offspring school performance (11).

The backdrop of conflicting practice, guidelines, and *ad hoc* translation of evidence to practice sees a clinical landscape that is confusing to women and practitioners alike. The dilemma for clinicians and pregnant women is founded in balancing the risks of i) causing iatrogenic short- and long-term harm associated with non-medically indicated IOL vs. ii) the risks of poorer perinatal outcomes with later or no intervention. The solution to be investigated in this study is a simple, cost-effective, and widely accessible approach that seeks to optimise and potentiate maternal physiology in late term to improve spontaneous labour rates, thereby reducing the need for IOL.

Induced by circadian cycles of light and darkness, maternal melatonin levels naturally peak at night under the influence of circadian control and trend upwards during pregnancy (16). Melatonin levels rise in response to falling daylight and peak between 2300 hrs and 0400 hrs (17). From 24 weeks’ gestation, the uterine muscles begin a diurnal pattern of contraction, two-thirds of which occur at night, under the influence of nocturnal melatonin levels (16). These contractions are largely imperceptible to the mother but increase in strength and frequency as the pregnancy progresses to late term. Recent *in vivo* observations and *ex vivo* findings demonstrate synergistic actions between melatonin and oxytocin demonstrating that melatonin sensitises myometrial cells to oxytocin, driving pulsatile contractions of the uterine muscles (18-20). Since most (72%) spontaneous labour occurs at night (21), which is when maternal serum melatonin levels peak (17), the synergistic action between melatonin and oxytocin is a natural and feasible link to test in this clinical trial.

Contemporary environmental and lifestyle factors are known to negatively impact upon endogenous melatonin production and release. Exposure to blue light through screens and mobile devices, sleep disturbance, shift work and changes to lifestyle factors such as diet and exercise, can all decrease melatonin synthesis and release (18, 22, 23). A clinical trial conducted in the United States examined the melatonin levels of late-term pregnant women who were exposed to short-wave light (known to inhibit endogenous melatonin levels). Investigators monitored uterine activity and melatonin levels before, during and after the intervention. Uterine activity and melatonin levels were reduced by more than 50% in those who were exposed to the short-wave light intervention. After intervention was withdrawn, serum melatonin levels restored to baseline and the uterine activity continued. These findings highlight the sensitivity of melatonin release to environmental triggers and the impact of melatonin on uterine contractility. Investigators concluded that exogenous melatonin may be a useful addition to the care of late term pregnancy, to counter the influence of environmental changes, or as an adjuvant to labour (18). In a recent observational study, this understanding was applied and further extended in a study conducted on women with late-term pregnancies. In Türkiye, melatonin levels were obtained from 362 women of varying gestations from term (40 weeks) to late term (42 weeks). Lower melatonin levels were highly predictive for prolonged pregnancy without labour. Higher melatonin levels were found in term and post-term women who laboured spontaneously. Investigators concluded that melatonin could be considered as a therapeutic agent to support effective labour (24). These studies point to the role melatonin may play in spontaneous onset of labour and uterine contractility and thus provide support for the use of melatonin in this trial.

## Rationale

In many OECD nations around the world, rates of induction of labour have increased sharply in the last decade (5, 6, 25). Despite escalated investment and medical intervention, rates of stillbirths, neonatal deaths and maternal mortality remain largely unchanged. Conversely, rates of some maternal and neonatal morbidity indicators have increased (9, 26). Despite its rising prevalence, induction of labour is not a benign intervention, associated with increased risk of caesarean section in nulliparous mothers, birth trauma and signals for potential longer term childhood impacts (2, 4, 9-12). Contemporary changes to human behaviour and environments that increase light exposure, can interfere with melatonin synthesis and release which may contribute to prolonged pregnancy (18, 22, 23). Melatonin supplementation is known to be safe in pregnancy (16, 27-30). Late term supplementation with melatonin may optimise maternal physiology and reduce the need for induction of labour by promoting the spontaneous onset of labour.

## Aim

The aim of this clinical trial is to determine if oral supplementation with 3 mg melatonin nightly from 39 weeks’ gestation in nulliparous women will reduce induction of labour rates.

## METHODS AND ANALYSIS

This protocol has been developed using an approved template for Clinical Trials, which is based on Therapeutic Goods Administration guidelines, SPIRIT guidelines and World Health Organization recommendations.

## Study Design

This study is a phase 3 placebo-controlled double-blind, randomised clinical trial. Trial design with nested sub-studies is shown in Fig. 1.

**Figure 1.**
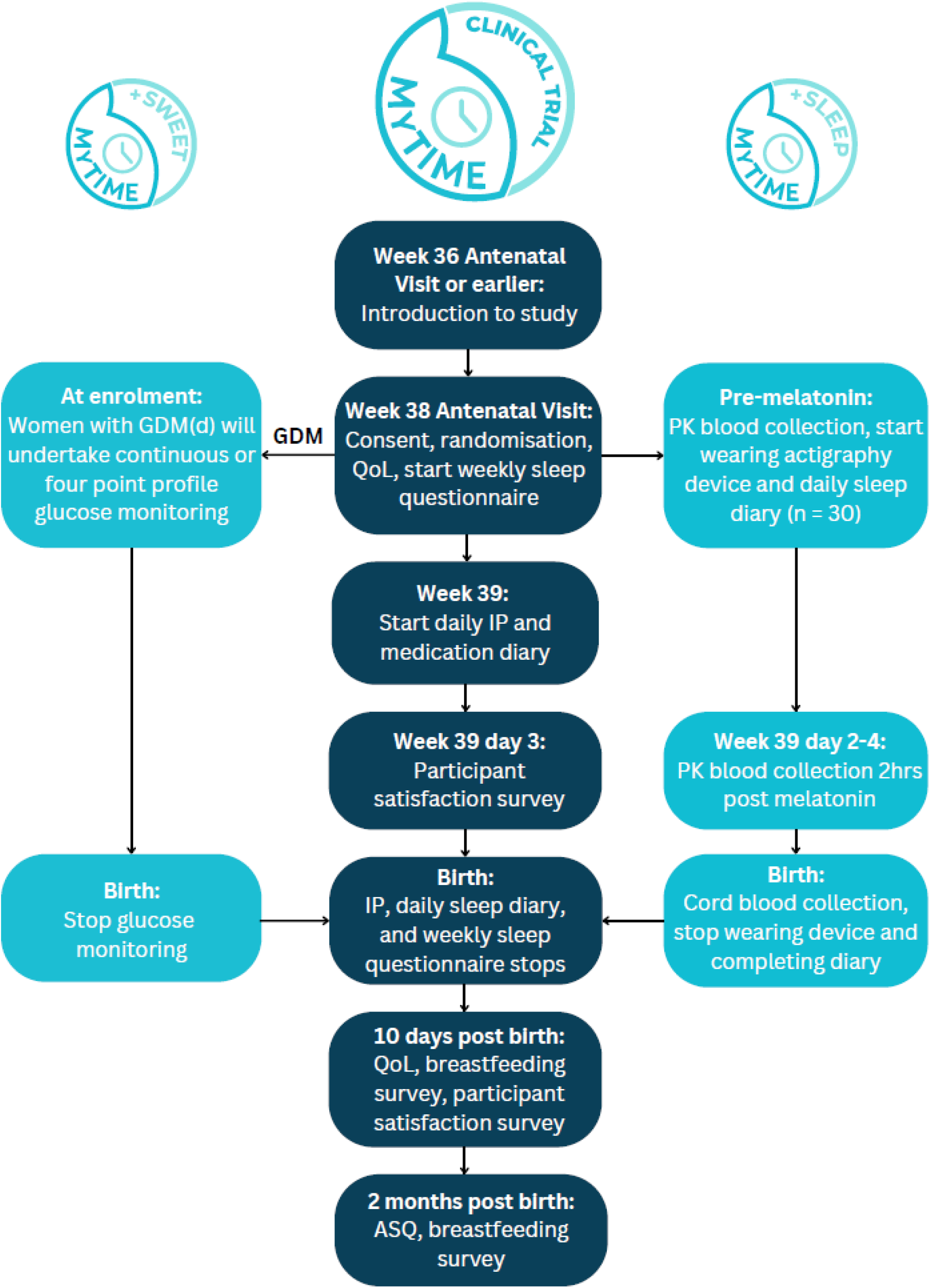
Trial design with nested sub-studies.

## Sub-studies

It is widely understood that melatonin plays an important role in supporting the onset and continuation of sleep (31). There is also emerging evidence that melatonin plays a role in regulating blood glucose levels (32, 33). Two sub-studies investigating impact on women’s sleep (MyTIME + Sleep) and blood glucose control (MyTIME + Sweet) are embedded within the trial design (Fig.1).

## Trial Timeline

The planned timeline is January 2024 – January 2027.

## Study Setting

The trial will be conducted within Women and Newborn Health Service (WNHS) the largest maternity service in Western Australia. The service cares for around 8000 women per year giving birth across the Perth metropolitan area as well as tertiary referral care for women from around Western Australia, the largest area health service in the world. Care is provided through a range of settings and models including: i) a quaternary/tertiary referral maternity hospital (King Edward Memorial Hospital -KEMH);ii)a secondary maternity service (Osborne Park Hospital); iii) primary birthing services offered through the state’s only stand-alone birth centre (Family Birth Centre); and iv) publicly funded homebirth program (Community Midwifery Program).

## Participants

Pregnant nulliparous women at 39 weeks’ gestation who meet inclusion criteria, booked to birth at one of the settings within the WNHS. Other inclusion and exclusion criteria are detailed in Table 1.

**Table 1.**
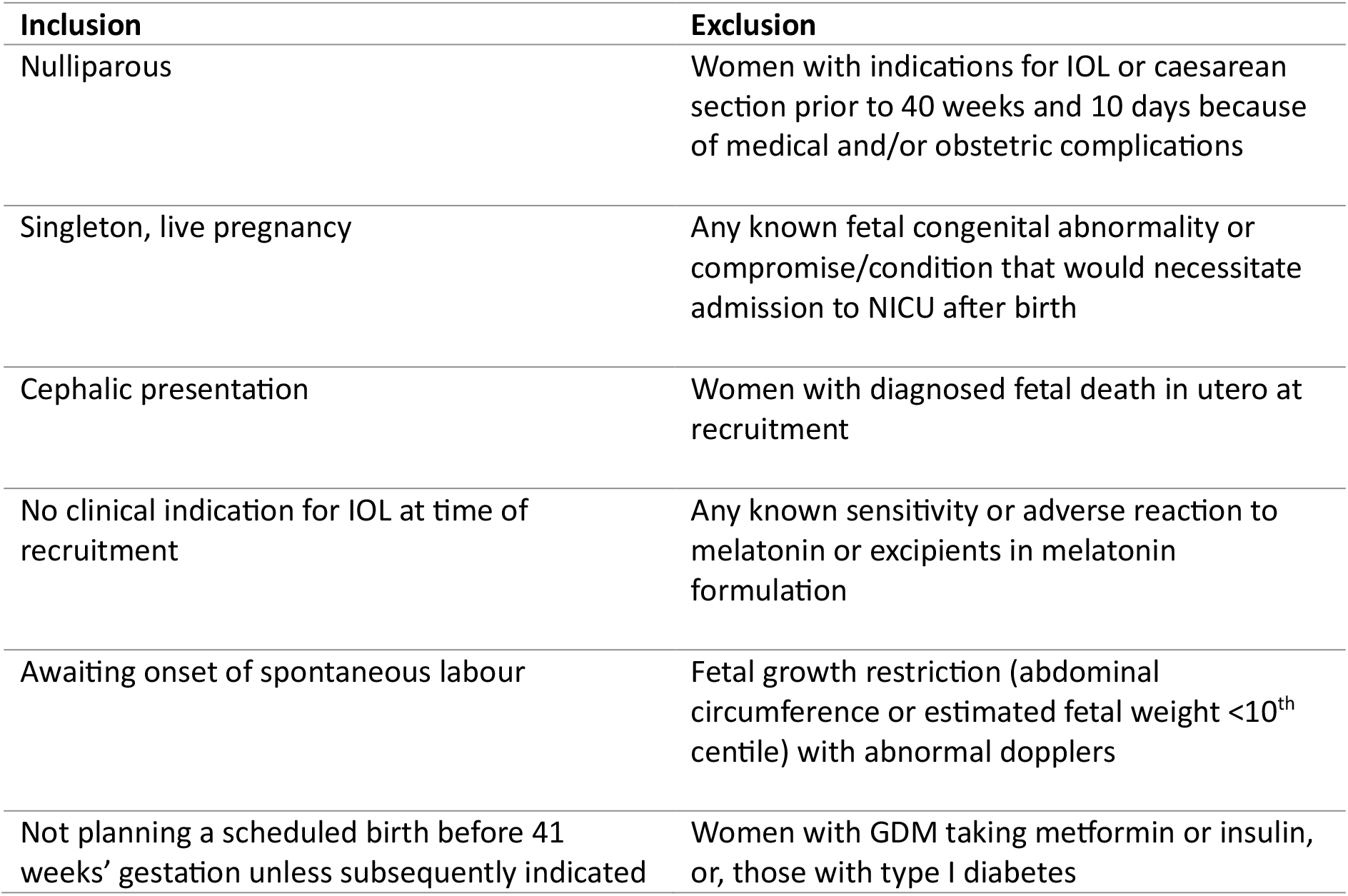
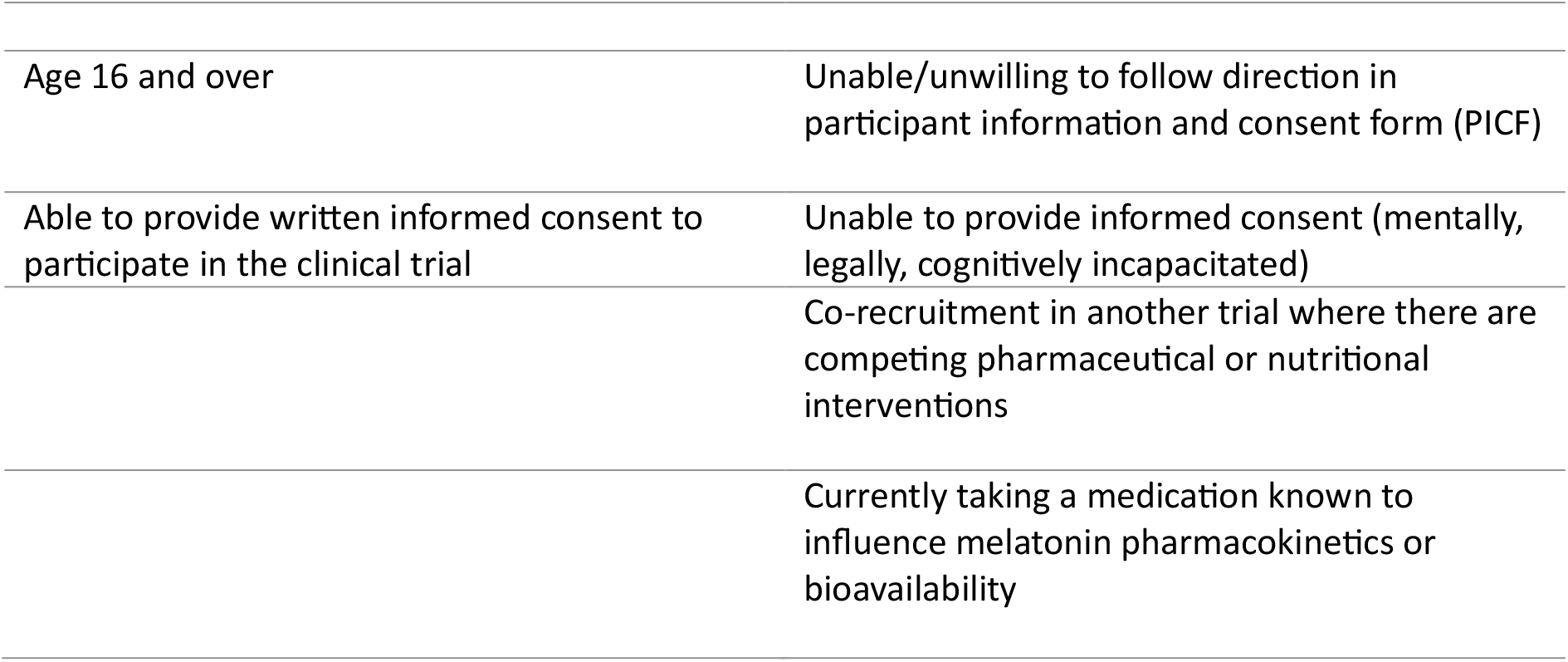
Inclusion and Exclusion Criteria.

Whilst not a specific criterion for inclusion, clarification is offered that women with GDM (not taking metformin or insulin) are able to be included in this trial. Women who require the aid of a language interpreter are able to be included in this trial.

## Recruitment and Informed Consent

Eligible potential participants will be approached at the various sites by a clinical trial midwife at their routine 36-week antenatal appointment who will provide them with information about the trial. If the potential participant is interested, the participant information form will be given to them to read and consider. Information will also be provided upon an eligible potential participant contacting the study team, if they hear about the trial from a recruitment poster or other source.

At the potential participant’s routine week 38 antenatal appointment, if they are willing to participate, they will provide written informed consent to the trial.

A subset of n=30 participants will be invited to participate in the MyTIME+Sleep sub-study (n=15 from each of the melatonin and placebo groups). Participants will be asked to give permission for a trial midwife to visit their home at night to collect pre/ post melatonin bloods and cord blood after birthing. Melatonin measurements are for ascertaining the efficacy of the intervention in elevating maternal circulating melatonin levels. Participants will also wear an actigraphy device (Axivity Model AX3) to provide objective sleep data.

All participants with diet controlled GDM will be asked to participate in the MyTIME+Sweet sub-study. It is anticipated that up to 150 women may be eligible to participate during the planned trial data collection period of two years. Sub study participation involves wearing a (trial supplied) continuous glucose monitor (CGM) for the trial duration. Those who do not wish to wear a CGM may continue in the trial with routine GDM care undertaking a four-point profile involving a finger prick blood glucose test four times a day.

## Randomisation and Blinding

A statistician will prepare the randomisation sequence for the trial, which will be implemented in the trial’s database. Participants will be randomised to melatonin or placebo in a 1:1 ratio. Randomisation will be stratified by model of care (continuity or non-continuity), GDM, and MyTIME+Sleep sub-study enrolment.

## Study Intervention

The study intervention is melatonin slow release (3 mg) encapsulated tablet administered orally in the evening. Both the melatonin and placebo will be compounded for the purpose of this double-blind trial, to ensure the tablets will be identical in appearance, weight, shape, and colour as to remain indistinguishable from each other.

At enrolment, participants will be automatically randomised and trial medication dispensed. Those enrolled in the MyTIME+Sweet sub study will be fitted with CGM to monitor pre- and intra-intervention blood glucose control. Those enrolled in MyTIME+Sleep sub study will wear an actigraphy device and arrange for pre and intra-intervention blood collection at night. On the night of the 39^th^ week, participants will receive a text message reminder to take trial medication in the evening, text reminders continue every evening until the participant indicates that baby has been born.

## Outcomes

The primary outcome is induction of labour after 39 weeks. Secondary outcomes include the interval between administration of trial medication and birth and a range of birth outcomes. Birth outcomes include onset and duration of labour; analgesia use during labour or birth; mode of birth; indication for mode of birth; estimated blood loss after birth; shoulder dystocia requiring at least one recorded manoeuvre; severe perineal trauma. A range of neonatal outcomes will also be considered including gestational age at birth; Apgar scores at 5 minutes; birth weight; admission to NICU within the first 24 hours of life; primary reason for admission, and length of stay if admitted; blood glucose levels for neonates of GDM mothers. Additional outcomes include mother and baby total length of stay; maternal blood glucose levels for GDM mothers before and during trial medication use; maternal biomarkers of inflammation and oxidative stress; maternal and cord blood plasma melatonin levels; breastfeeding on discharge, at 10 days, and at 2 months; perinatal mortality; maternal trial participation satisfaction; quality of life adjusted years (QALY) pre and post-trial; sleep duration via self-report and (n=30) via actigraphy; sleep quality rating (self-reported); Ages and Stages Questionnaire ASQ-2M assessing child developmental outcomes at 2 months of age; cost effectiveness of melatonin compared with standard care.

A pre-planned analysis of the primary and secondary outcomes stratified by BMI and Model of Care will also be performed.

## Maternal and Pregnancy Characteristics

The following characteristics will be collected and may be considered in modelling. Maternal date of birth, weight and height at time of booking; Aboriginal or Torres Strait Islander (Australian Indigenous people); maternal self-reported ethnicity, country of birth, language spoken at home; length of time in Australia; Body Mass Index at booking; medical conditions including diagnosis of anxiety or depression; gravidity and parity; medications in pregnancy; substance use in pregnancy; sexually transmitted infections in pregnancy; alcohol intake in pregnancy; tobacco or vaping in pregnancy; Group B Streptococcus screening results; Edinburgh Postnatal Depression Scale scores; Family Domestic Violence Screening; previous blood donation; number of antenatal visits; setting of labour and birth; antenatal complementary therapies discussed with care providers and recorded in the perinatal database, and special child health referral.

## Sampling and Data Collection

Data will be obtained from various sources: STORK (the clinical perinatal database used by maternity services within WA public health services), maternal medical record (via digital record or paper file), directly from the participant (in person or via questionnaire), directly from clinicians, blood tests, and wearable devices. NEObase (the neonatal admissions database at KEMH) will be accessed only if required for neonatal safety reporting.

Participant data collected will be entered into REDCap for analysis.

A total sample size of 530 women (∼265 per group) will attain 80% power to detect this clinically relevant reduction of induction rate (30%) in the melatonin group (odds ratio of 0.57) while using logistic regression analysis with adjustment for the stratification factors and other relevant covariates with an r-squared of 0.025 at alpha=0.025. This sample size is also inflated to account for a 10% loss to follow-up (Power and Sample Size Program for Windows, version 2019).

## Statistical Analysis

Data will be analysed on intention-to-treat basis. Binomial and logistic regression analyses will be performed on primary endpoint and other binary outcomes. Linear and/or Cox proportional hazards regression will be used to examine group differences between the continuous and time to event outcomes. Melatonin adherence will be assessed and, if applicable, supplementary analyses on the adherent subgroup and per treatment received will also be performed. All hypothesis tests will be two-sided with alpha=0.05. Data analyses will be performed using STATA statistical software (version 16). A single blinded interim safety analysis will be conducted when 50% of the participants (N=265) have been recruited.

A within trial cost-effectiveness analysis will be conducted to compare differences in costs and QALYs of women receiving melatonin supplementation compared to those receiving standard care. Costs and QALYs will be compared using generalised linear models. A modelled cost-effectiveness analysis will then be conducted to assess cost-effectiveness and budget impact with population level implementation and projecting to a five-year time horizon to estimate long-term cost effectiveness.

## Safety Events

Monitoring, assessment, and reporting of adverse events within the trial will occur as per the National Health and Medical Research Council Guidance: Safety monitoring and reporting in clinical trials involving therapeutic goods (2016). All safety events will be assessed regardless of causal relationship. Identification of safety events may occur via alert from the participant and/or observed by the researcher and/or clinical staff, and/or identified in the course of other trial related procedures.

Oral melatonin is known to be safe for pregnant women and babies. There are no anticipated risks from taking oral melatonin during pregnancy, and published accounts of clinical studies in pregnant populations do not report serious adverse reactions or safety concerns. Recent cohort studies researching melatonin supplementation in the general population have reported encouraging pharmacotherapeutic findings, including regulation of hypertension, protection against maternal and neonatal oxidative stress, neuro-regulation, and neuro-protection. However, as with any pharmaceutical, there is risk of a participant experiencing an unexpected, previously unknown adverse reaction or hypersensitivity to melatonin. Safety events will be reported from randomisation up until 24 hours after the final administration of melatonin/placebo. This reporting period was selected as it more than covers 5 half-lives of the study drug.

All safety events assessed as possibly, probably, or definitely related to the trial medication and all serious safety events will be followed up to resolution, or until they are assessed as stabilised but unlikely to resolve.

The admission of a baby to the NICU within 24 hours after the final maternal administration of melatonin/placebo will be reported as a serious adverse event. Prolonged hospitalisation for a reason unrelated to trial participation, such as social reasons or planned hospitalisation for the purpose of birth will not be reported. However, if birth complications result in prolonged hospitalisation, this will be reported.

## Data and Safety Monitoring Committee (DSMC)

A DSMC will be established as melatonin is not currently approved for use in pregnancy, or to promote onset of spontaneous labour. The DMSC will review study progress, recruitment, and adverse events. The committee will be comprised of an independent obstetrician, neonatologist, midwife, biostatistician, and pharmacist and will consider and suggest any changes to the protocol which may be recommended.

Data will be presented without linkage to any participant identifiers. Data will also remain blinded to the DSMC members. If a safety signal is apparent, the DSMC will request for data to be unblinded. Data will be provided to the DSMC 6 monthly, or at request of the committee, the Sponsor (Curtin University), or the HREC. An interim safety analysis will also be carried out at 50% recruitment, and results will be provided to the DSMC for review. This analysis will compare various perinatal outcomes for MyTIME recruits compared to the wider WNHS primiparous population.

The WNHS Human Research Ethics Committee (HREC), WNHS Research Governance Office (RGO), and the Sponsor, will be notified of DSMC findings at each safety assessment.

## Trial Discontinuation

The trial may prematurely, permanently, or temporarily cease recruitment if the PI, DSMC, or the Sponsor, believe there are issues pertaining to participant welfare and safety; a serious breach of trial protocol; a recommendation from the DSMC that the trial should cease or be re-evaluated.

If the trial is ceased prematurely, the Sponsor, WNHS HREC, and WNHS RGO will be immediately informed.

## Unblinding

The trial may be unblinded in the following circumstances: to make clinical treatment decisions when an unexpected serious adverse event occurs, and the intervention must be known; at the request of the DSMC; at the conclusion of the trial to determine intervention effectiveness as per study protocol

## Public Involvement

Considerable engagement with consumers has informed the project conceptualisation and development. Our pre-trial consumer data indicate high acceptability and strong maternal demand for this proposed innovative trial, with 90% of (n= 172) women surveyed indicating they would be interested in participating if this trial were available to them (34).

We have a consumer representative as a member on our trial team who was involved in the successful grant application and has contributed to the review and design of this study at each stage. Consumer involvement is embedded at every stage of this trial and will continue through to knowledge translation. The mutual investment of consumer representation in the conduct of this RCT supports knowledge translation and capacity to influence maternity policy.

## Ethics and Dissemination

The study is approved by the WNHS HREC (RGS0000006283) and will be carried out in accordance with the approval conditions of the HREC. The trial will also be carried out in accordance with all applicable guidelines set out by the NHMRC.

Trial findings will be disseminated through conference presentations and peer reviewed publications. We will also co-design a consumer facing infographic containing key data from this trial to be disseminated via the trial consumer representative with established national and international networks. We will also ensure opportunities for the consumer representative to join communication of trial findings to clinicians through established professional networks.

## Discussion

Induction of labour has increased without clear evidence of improvement in perinatal outcomes for those mothers who do not have a medical indication for induction (1-6). It is plausible that contemporary environmental and lifestyle factors may have an inhibitory impact on synthesis and release of melatonin (18, 22, 23), a hormone shown to be involved in spontaneous labour (17, 24). Melatonin supplementation may potentiate late term maternal physiology and reduce the need for induction of labour in women who do not have a medical indication for intervention. If so, this affordable, accessible, off-patent medication would constitute an acceptable alternative to induction of labour which carries risk of iatrogenic harm. Melatonin is known to be safe in pregnancy (16, 27-30) and offers a range of potential health benefits and may play a role in supporting spontaneous labour (17, 24).

## Trial Status

The trial commenced recruitment in January 2024. Two years has been allocated for recruitment based on service data.

## Data Availability

All data produced in the present study are available upon reasonable request to the authors

## Authors’ Contributions

A/Prof Zoe Bradfield conceptualised the trial.

A/Prof Zoe Bradfield, Prof Jeffrey Keelan, Dr Scott White, A/Prof Mary Sharp, Dr Miranda Davies-Tuck, A/Prof Jane Warland, Dr Lesley Kuliukas, Prof Dorota Doherty, Prof Emily Callander, Kylie Ekin, Monique Rose, and Amber Pettitt designed and wrote the trial protocol.

## References

1. Acsqhc, Aihw. The Fourth Australian Atlas of Healthcare Variation. Sydney: ACSQHC AIHW; 2021.

2. Adler K, Rahkonen L, Kruit H. Maternal childbirth experience in induced and spontaneous labour measured in a visual analog scale and the factors influencing it; a two-year cohort study. BMC Pregnancy and Childbirth. 2020;20(1):415.

3. Butler SE, Wallace EM, Bisits A, Selvaratnam RJ, Davey M-A. Induction of labor and cesarean birth in lower-risk nulliparous women at term: A retrospective cohort study. Birth. 2024;n/a(n/a).

4. Dahlen HG, Thornton C, Downe S, De Jonge A, Seijmonsbergen-Schermers A, Tracy S, et al. Intrapartum interventions and outcomes for women and children following induction of labour at term in uncomplicated pregnancies: a 16-year population-based linked data study. BMJ Open. 2021;11(6):e047040.

5. Haavaldsen C, Morken N-H, Saugstad OD, Eskild A. Is the increasing prevalence of labor induction accompanied by changes in pregnancy outcomes? An observational study of all singleton births at gestational weeks 37–42 in Norway during 1999–2019. Acta Obstetricia et Gynecologica Scandinavica. 2023;102(2):158–73.

6. McCarthy CM, Meaney S, McCarthy M, Conners N, Russell N. Induction of labor: reviewing the past to improve the future. AJOG Global Reports. 2022;2(4):100099.

7. White SW, & Newnham, J. P. Is it possible to safely prevent late preterm and early term births? Seminars in fetal & neonatal medicine. 2019;24(1), 33–36.

8. Yin W, Döring N, Persson MSM, Persson M, Tedroff K, Ådén U, et al. Gestational age and risk of intellectual disability: a population-based cohort study. Arch Dis Child. 2022:archdischild-2021-323308.

9. Aihw. Australia’s Mothers and Babies. Canberra: Australian Institute of Health and Welfare; 2022.

10. Health AIo, Welfare. Maternal deaths. Canberra: AIHW; 2023.

11. Berger BO, Jeffers NK, Wolfson C, Gemmill A. Role of Maternal Age in Increasing Severe Maternal Morbidity Rates in the United States. Obstet Gynecol. 2023;142(2):371–80.

12. Lin L, Ren LW, Li XY, Sun W, Chen YH, Chen JS, et al. Evaluation of the etiology and risk factors for maternal sepsis: A single center study in Guangzhou, China. World J Clin Cases. 2021;9(26):7704–16.

13. WHO Guidelines Approved by the Guidelines Review Committee. WHO recommendations: Induction of labour at or beyond term. Geneva: World Health Organization © World Health Organization 2018.; 2018.

14. Declercq E, Belanoff C, Iverson R. Maternal perceptions of the experience of attempted labor induction and medically elective inductions: analysis of survey results from listening to mothers in California. BMC Pregnancy and Childbirth. 2020;20(1).

15. Nagle U, Naughton S, Ayers S, Cooley S, Duffy RM, Dikmen-Yildiz P. A survey of perceived traumatic birth experiences in an Irish maternity sample – prevalence, risk factors and follow up. Midwifery. 2022;113:103419.

16. McCarthy R, Jungheim ES, Fay JC, Bates K, Herzog ED, England SK. Riding the Rhythm of Melatonin Through Pregnancy to Deliver on Time. Front Endocrinol (Lausanne). 2019;10:616-.

17. Olcese J, Lozier S, Paradise C. Melatonin and the Circadian Timing of Human Parturition. Reproductive Sciences. 2013;20(2):168–74.

18. Rahman SA, Bibbo C, Olcese J, Czeisler CA, Robinson JN, Klerman EB. Relationship between endogenous melatonin concentrations and uterine contractions in late third trimester of human pregnancy. Journal of Pineal Research. 2019;66(4):e12566.

19. Sharkey JT, Puttaramu R, Word RA, Olcese J. Melatonin Synergizes with Oxytocin to Enhance Contractility of Human Myometrial Smooth Muscle Cells. The Journal of Clinical Endocrinology & Metabolism. 2009;94(2):421–7.

20. Sharkey JT, Cable C, Olcese J. Melatonin Sensitizes Human Myometrial Cells to Oxytocin in a Protein Kinase Cα/Extracellular-Signal Regulated Kinase-Dependent Manner. The Journal of Clinical Endocrinology & Metabolism. 2010;95(6):2902–8.

21. Martin P, Cortina-Borja M, Newburn M, Harper G, Gibson R, Dodwell M, et al. Timing of singleton births by onset of labour and mode of birth in NHS maternity units in England, 2005–2014: A study of linked birth registration, birth notification, and hospital episode data. PLOS ONE. 2018;13(6):e0198183.

22. Mitsui K, Saeki K, Tone N, Suzuki S, Takamiya S, Tai Y, et al. Short-wavelength light exposure at night and sleep disturbances accompanied by decreased melatonin secretion in real-life settings: a cross-sectional study of the HEIJO-KYO cohort. Sleep Med. 2022;90:192–8.

23. Rahman SA, Wright KP, Lockley SW, Czeisler CA, Gronfier C. Characterizing the temporal Dynamics of Melatonin and Cortisol Changes in Response to Nocturnal Light Exposure. Scientific Reports. 2019;9(1).

24. Yurtcu N, Caliskan C, Celik S. Serum Melatonin as a Biomarker for Assessment of Late-term and Postterm Pregnancies in Women without Spontaneous Onset of Labor. Z Geburtshilfe Neonatology. 2021;6:6.

25. Swift EM, Gunnarsdottir J, Zoega H, Bjarnadottir RI, Steingrimsdottir T, Einarsdottir K. Trends in labor induction indications: A 20-year population-based study. Acta Obstet Gynecol Scand. 2022;101(12):1422–30.

26. Flood M, McDonald SJ, Pollock W, Cullinane F, Davey M-A. Incidence, trends and severity of primary postpartum haemorrhage in Australia: A population-based study using Victorian Perinatal Data Collection data for 764 244 births. Australian and New Zealand Journal of Obstetrics and Gynaecology. 2019;59(2):228–34.

27. Carloni S, Proietti F, Rocchi M, Longini M, Marseglia L, D’Angelo G, et al. Melatonin Pharmacokinetics Following Oral Administration in Preterm Neonates. Molecules. 2017;22(12):2115.

28. Tarocco A, Caroccia N, Morciano G, Wieckowski MR, Ancora G, Garani G, et al. Melatonin as a master regulator of cell death and inflammation: molecular mechanisms and clinical implications for newborn care. Cell Death & Disease. 2019;10(4).

29. Vine T, Brown GM, Frey BN. Melatonin use during pregnancy and lactation: A scoping review of human studies. Brazilian Journal of Psychiatry. 2022;44(3):342–8.

30. Wei S, Smits MG, Tang X, Kuang L, Meng H, Ni S, et al. Efficacy and safety of melatonin for sleep onset insomnia in children and adolescents: a meta-analysis of randomized controlled trials. Sleep Med. 2020;68:1–8.

31. Costello RB, Lentino CV, Boyd CC, O’Connell ML, Crawford CC, Sprengel ML, et al. The effectiveness of melatonin for promoting healthy sleep: a rapid evidence assessment of the literature. Nutr J. 2014;13:106.

32. Patel R, Parmar N, Pramanik Palit S, Rathwa N, Ramachandran AV, Begum R. Diabetes mellitus and melatonin: Where are we? Biochimie. 2022;202:2–14.

33. Pourhanifeh MH, Hosseinzadeh A, Dehdashtian E, Hemati K, Mehrzadi S. Melatonin: new insights on its therapeutic properties in diabetic complications. Diabetol Metab Syndr. 2020;12:30.

34. PSANZ 2023 - Oral. Journal of Paediatrics and Child Health. 2023;59(S1):4–54.

